# Staging and typing of chest CT images: A quantitative analysis based on an ambispective observational cohort study of 125 patients with COVID-19 in Xiangyang, China

**DOI:** 10.1101/2020.10.25.20219253

**Authors:** Guoxin Huang, Yong Wang, Xiaxia Wu, Gaojing Qu, Junwen Chen, Hui Yu, Meiling Zhang, Lisha Wang, Jinwei Ai, Haoming Zhu, Lei Chen, Bin Pei

## Abstract

**Background:** The stage of CT images was rarely studied and the relationship between the severity of Coronavirus Disease 2019 (COVID-19) and CT images has not been studied based on systematic quantitative analysis currently.

**Purpose:** To investigate the staging duration and classification of CT images of patients with COVID-19 based on quantitative analysis.

**Materials and Methods:** This is an ambispective observational cohort study based on 125 patients with COVID-19 from Jan 23 to Feb 28, 2020. The stage of CT and pulmonary lesion size were quantitatively analyzed. The categorical regression analysis based on optimal scale (CATREG) was performed to evaluate the association of CT score, age, and gender with the clinical type.

**Results:** The CT images of 125 patients with COVID-19 (50.13 ± 16.91 years, 66 women) were analyzed in this study. Except for pre-early stage, the duration of early, progression-consolidation, and dissipation stage of CT images was 3.40 ± 2.31, 10.07 ± 4.91, and 20.60 ± 7.64 days, respectively. The median CT score was 5.00 (2.00-8.50) during the first 30 days, which reached a peak on the 11^th^ day. Significant differences were found between the median CT scores of different clinical types (P<0.05). Besides, the age was correlated with the clinical type (P<0.001), the CT scores of 0.00-11.50, 11.50-16.00, and 16.00-20.00 were separately correlated with the moderate, severe, and critical type with the output accuracy 69.60%.

**Conclusion:** The four-stage staging method based on quantitative analysis is consistent with the change rules of staging features and COVID-19. Quantitative study by scoring pulmonary lesion sizes accurately revealed the evolvement of pulmonary lesions and differences between different clinical types.

**Summary:** Quantitative study of the stage duration and classification of chest CT images can objectively reveal the relationship between Coronavirus Disease 2019 (COVID-19) and chest CT images.

**Key Results:** 1. A four-stage staging method was proposed. Except for pre-early stage, the duration of early, progression-consolidation, and dissipation stage of CT images was 3.40 ± 2.31, 10.07 ± 4.91, and 20.60 ± 7.64 days, respectively.

2. The severer the disease, the higher the median CT scores and their peak value.

3. The CT scores of 0.00-11.50, 11.50-16.00, and 16.00-20.00 were separately correlated with the moderate, severe, and critical type.

## Introduction

The Coronavirus Disease 2019 (COVID-19), caused by SARS-CoV-2 infection, has spread around the world and become a global pandemic declared by WHO since March 11, 2020^[1, 2]^. Chest CT examination has revealed pathological changes in the lungs of patients with COVID-19, most of which showed multiple small patchy shadow and changes in the interstitial lungs especially peripheral region at the early stage, and then developed into multiple bilateral ground-glass opacity (GGO)^[3]^. Because of its definite and credible nature of images, CT examination has been highly recommended in the guideline for COVID-19 diagnosis and treatment^[4]^.

Several in-depth studies have been performed to investigate the application of CT in COVID-19 diagnosis^[5-7]^. The features of CT images were different at the distinct stages of the disease course^[8, 9]^. The relevant guidelines have put forward the durations of CT stages^[10, 11]^. Pan et al^[12]^. and Li et al^[13]^. specifically staged the CT images based on quantitative analysis.

Currently, there are two methods for quantitatively studying CT images of pulmonary lesions. The first one divided each lateral lungs into the upper, middle, and lower regions, separately rated 25, 50, 75, and 100% lesion in each region as 1, 2, 3, and 4 points; and rated the lesions nature with CT attenuation, GGO, and consolidation as 1, 2, and 3 points, respectively^[14-16]^. However, this method involved the nature of pulmonary lesions during scoring, and could not accurately reveal the pulmonary lesion sizes. The other method calculated the percentage of lesion area in the whole lung by software^[17, 18]^, which cannot be carried out in most hospitals currently. Besides, the relationship between CT scores and the severity of disease has not been analyzed in the above methods.

In this study, we established a cohort of 125 patients with COVID-19 in Xiangyang, Hubei, China, described the staging duration of their CT images, and put forward a four-stage staging method based on the time of symptom onset and appearance time of CT features at each stage. Moreover, a quantitative analysis for classifying these CT images was performed using a methodology developed by Wu et al. based on anatomy of the lungs^[19]^. The size and change of pulmonary lesions were assessed and scored, and the relationship between the scores and the severity of the disease was evaluated. Our findings may be useful in diagnosis and treatment of COVID-19.

## Materials and Methods

### Study design

This study is an ambispective observational cohort study.

### Patients

This cohort was established on Feb 9, 2020. The suspected and confirmed patients with COVID-19 in this study were admitted to the Xiangyang No.1 People’s Hospital affiliated hospital of Hubei University of Medicine according to the Diagnosis and Treatment Protocol for Novel Coronavirus Pneumonia^[3]^. The retrospective data were traced back to Jan 23, 2020. The follow-up was carried out until Mar 28, 2020. The study was approved by the ethics review board at Xiangyang No.1 People’s Hospital (No. 2020GCP012). Informed consent from patients has been exempted, which neither involves the personal privacy of patients nor incurs greater than the minimal risk. Chinese clinical trial registry No.: ChiCTR2000031088. The severity of the patients was classified from mild to critical type and then cross-checked by two respiratory physicians. The inconsistency was resolved by a third party.

### Data collection

The clinical data of the patients with COVID-19 were collected independently from the hospital information system and four-cross-checked, including hospitalization registry numbers, genders, ages, symptom onset time, all symptoms, inception dates of critical type, chest CT images, laboratory test results, and outcomes.

### CT scanning

Chest CT scanning from apex to base of the lungs was performed on an Aquilion TSX101A CT scanner (Toshiba, Japan) or an Aquilion PRIME CT scanner (Toshiba, Japan) with a 64-row or 80-row detector, respectively. Scanning parameters for the lungs: tube voltage 120kV, Matrix size 256×256, FOV 379.7 (L). Helical pitch was separately set at Fast (1.484 / HP 95) mm and Fast (1.388 / HP 111.0) mm in the 64-row and 80-row CT.

### Staging of chest CT images of patients with COVID-19

Two senior radiologists independently analyzed all CT images, described the features of pulmonary manifestations in each CT image, and matched them with the staging criteria in the Guidelines for Imaging Diagnosis of novel Coronavirus Infected Pneumonia^[20]^. The duration from symptom onset to the CT examining date was calculated for all CT images. The mean duration and its standard deviation at each stage were calculated and cross-checked. The inconsistency was resolved by a third party.

### Quantitative analysis of pulmonary lesion sizes and their temporal changes

Two senior radiologists independently scored all pulmonary lesions in each CT images according to the method proposed by Wu et al.^[19]^. Regardless of the nature of pulmonary lesions, the largest lesion exceeded half of the segment in the axial thin-section CT image was scored 1 point, and the largest lesion less than half of the segment was scored 0.5 point. Every five days starting from the symptom onset day were designated as T1, T2, …, Tn, where T1 was the first five-day. The temporal changes of the median CT scores were calculated from symptom onset to the 30^th^ day, and separately illustrated in terms of per day and per five days. The pulmonary lesion sizes and their temporal changes of the whole cohort and different clinical types were separately quantitatively analyzed and compared.

### Categorical regression analysis

The categorical regression analysis with optimal scale (CATREG) was performed by setting the clinical type as dependent variable, setting the maximum CT score in the first 15 days, age, and gender as independent variables, and setting gender as nominal variable.

### The outcome measures

The outcome measures included the general information, clinical types, feature of manifestations in CT images and their appearance time at each stages and those for the specific clinical types, scores of pulmonary lesions in CT images, the medians and their temporal changes of CT scores in terms of per day and per five days for the whole cohort and for the different clinical types, outcomes, and CATREG model.

### Statistical analysis

All data were analyzed using SPSS20.0. Binary/dichotomous data were described by counts or percentages. Mean (M) and standard deviation (SD) were calculated to analyze the continuous data with normal distribution. Median and interquartile range (IQR) were calculated to analyze the continuous data with abnormal distribution. Chi square test was used for enumeration data; T-test and nonparametric test were used for continuous data to analyze statistical differences between groups. P<0.05 indicated a significant difference.

## Results

### General information

There were 400 patients with negative result in pharyngeal swab nucleic acid test and 142 patients with positive result admitted to our hospital until February 28, 2020. Among the positive nucleic-acid-test patients, the data of 9 patients transferred from other hospitals could not be traced back, 2 critical patients only had X-ray examination, 4 mild patients did not exhibit pneumonia manifestation, and 2 infant patients had different CT manifestations. Therefore, the CT images of 125 patients (50.13 ± 16.91 years, 66 women) were finally investigated in the following study, among which 88, 21, and 16 patients belonged to the moderate, severe, and critical type, respectively. The average duration from symptom onset to transforming into critical type was 12.50 ± 5.73 days.

### Features of pulmonary manifestations and staging of CT images

There were no pre-early-stage CT images in this cohort. As shown in Table 1 and Figure 1, the duration of early stage was 3.40 ± 2.31 days and characterized by that the lesions were localized and exhibited as single or multiple patchy or nodular GGO, mostly distributed under the pleura, mainly in sub-segments or segments. The duration of progression stage was 9.04 ± 4.51 days and characterized by that the density of some lesions unevenly increased, multiple lobes mainly the lower lobes were involved where there were air bronchogram and vascular thickening. High resolution CT showed fine grid-like changes. The duration of consolidation stage was 11.01 ± 5.09 days and characterized by that consolidation and cord shadow increased, GGO coexisted with patchy consolidation and cord-like interstitial thickening opacity. In a few cases, whole lungs presented a “white lungs” manifestation. The duration of dissipation stage was 20.60 ± 7.64 days and characterized by that complete absorption of lesions or patchy consolidation, and less absorption of strip shadows. Since the progression and consolidation stages were identified in the same CT images in 31.84% (57/179) of the examinations, we combined these two stages as the progression-consolidation stage with duration of 10.07 ± 4.91 days.

**Table 1:** The durations of different CT stages in the whole cohort and different clinical types.

| **Stages** | **Early**<br>**(day)** | **Progression**<br>**(day)** | **Consolidation**<br>**(day)** | **Dissipation**<br>**(day)** | **Progression-**<br>**Consolidation**<br>**(day)** |
| --- | --- | --- | --- | --- | --- |
| **Whole cohort** |  |  |  |  |  |
| Mean | 3.40 | 9.04 | 11.01 | 20.60 | 10.07 |
| SD | 2.31 | 4.51 | 5.09 | 7.64 | 4.91 |
| Sets of images (N) | 86 | 85 | 94 | 101 | 179 |
| **Moderate** |  |  |  |  |  |
| Mean | 3.46 | 8.75 | 10.21 | 18.04 | 9.52 |
| SD | 2.33 | 3.67 | 3.72 | 5.05 | 3.75 |
| Sets of images (N) | 61 | 56 | 63 | 76 | 119 |
| P1 | 0.755 | 0.4061 | 0.2404 | 0 | 0.6979 |
| **Severe** |  |  |  |  |  |
| Mean | 3.23 | 7.94 | 11.50 | 27.42 | 9.82 |
| SD | 2.65 | 3.15 | 5.68 | 8.61 | 4.94 |
| Sets of images (N) | 13 | 18 | 20 | 19 | 38 |
| P2 | 0.984 | 0.0501 | 0.2175 | 0.3393 | 0.0339 |
| **Critical** |  |  |  |  |  |
| Mean | 3.25 | 12.27 | 14.73 | 31.50 | 13.50 |
| SD | 2.01 | 8.08 | 8.58 | 9.97 | 8.23 |
| Sets of images (N) | 12 | 11 | 11 | 6 | 22 |
| P3 | 0.773 | 0.024 | 0.004 | 0.000 | 0.000 |

P1: moderate type vs. severe type; P2: severe type vs. critical type; P3: moderate type vs. critical type

**Figure 1:**
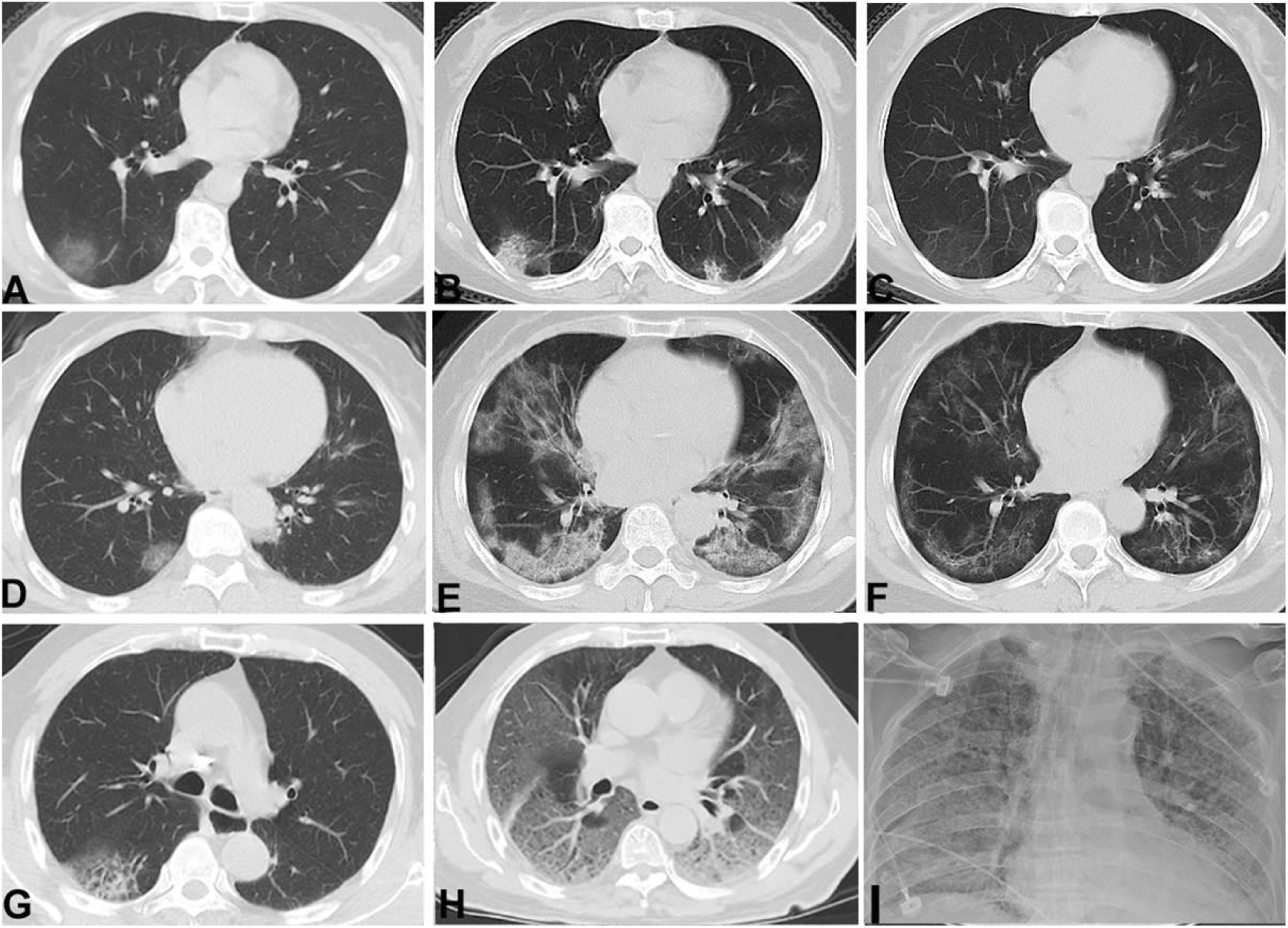
Typical CT images of different stages in different clinical types. A-C, The moderate type(female,58 years). (A) The early stage, scored 6 points, localized GGO under pleura in the lower lobes of the right and left lungs. (B) The progression-consolidation stage, scored 6 points, the lesions were increased in number and attenuation with consolidation and fibrosis in the lower lobes of the right and left lungs. (C) The dissipation stage, scored 4 points, the lesions were decreased in attenuation and mostly absorbed in the lower lobes of the right and left lungs. D-F, The severe type(female,54years). (D) The early stage, scored 3 points, multiple patchy GGO under pleura in the right and left lungs. (E) The progression-consolidation stage, scored 18 points, the lesions were increased in range and attenuation with partial consolidation and fibrosis in the right and left lungs. (F) The dissipation stage, scored 15.5 points, the lesions were decreased in range and attenuation in the right and left lungs. G-I, The critical type(male,75years). (G) The early stage, scored 1 point, patchy denser opacity with grid in it in the dorsal segment of the lower lobe of the right lungs. (H) The progression stage, 20 points, the number of the ground-glass denser opacity was significantly increased and merged and presented as “white lungs”. (I) Diffuse distribution of flake-like ground glass opacities with fuzzy boundaries in the right and left lungs. The participant was unable to receive CT examination but a digital Radiography examination.

### Duration of each CT stage in different clinical types

As shown in Table 1, for the moderate type, the durations of early, progression, consolidation, dissipation stage, and the combined progression-consolidation stage were 3.46 ± 2.33, 8.75 ± 3.67, 10.21 ± 3.72, 18.04 ± 5.05, and 9.52 ± 3.75 days, respectively. For the severe type, the durations of the corresponding stages were 3.23 ± 2.65, 7.94 ± 3.15, 11.50 ± 5.68, 27.42 ± 8.61, and 9.82 ± 4.94 days, respectively.

For the critical type, the durations of the corresponding stages were 3.25 ± 2.01, 12.27 ± 8.08, 14.73 ± 8.58, 31.50 ± 9.97, and 13.50 ± 8.23 days, respectively. The duration of dissipation stage in severe type was significantly longer than those in moderate type; the duration of combined progression-consolidation stage in critical type was significantly longer than those in severe type; the durations of the whole stages except the early stage in critical type was significantly longer than those in moderate type (P<0.05).

### Quantitative analysis of pulmonary lesion sizes and their temporal changes

The median CT score of the 125 patients was 5.00 (2.00-8.50) during the first 30 days (Table 2), which was 2.00 on the 1^st^ day and reached a peak of 8.25 on the 11^th^ day. The temporal change trends of median CT scores in terms of per day and per five days were separately illustrated in Figure 2A and Figure 2B.

**Table 2:**
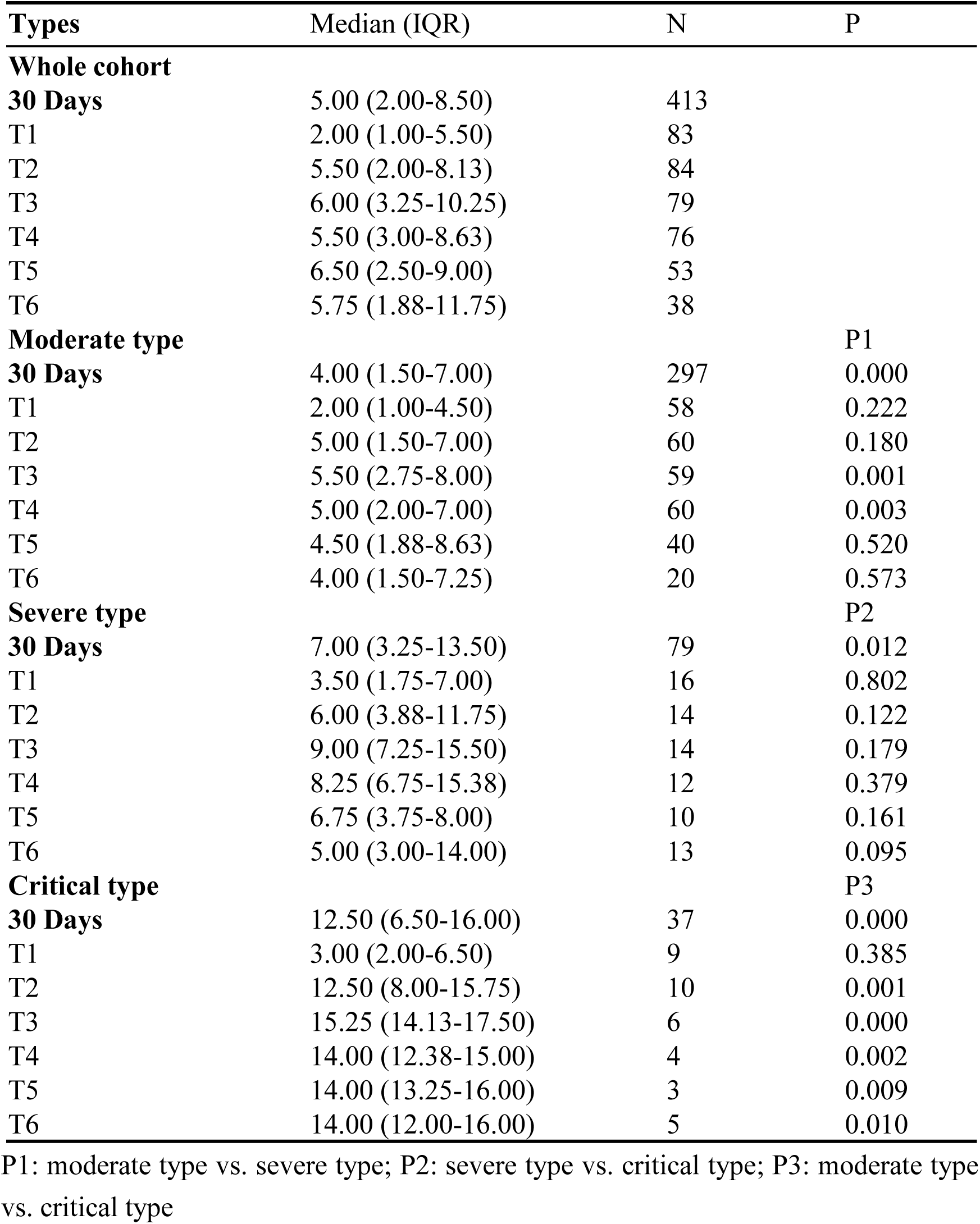
The median and IQR scores of pulmonary lesion sizes and their temporal changes in the whole cohort and different clinical types.

| Types | Median (IQR) | N | P |
| --- | --- | --- | --- |
| **Whole cohort** |  |  |  |
| **30 Days** | 5.00 (2.00-8.50) | 413 |  |
| T1 | 2.00 (1.00-5.50) | 83 |  |
| T2 | 5.50 (2.00-8.13) | 84 |  |
| T3 | 6.00 (3.25-10.25) | 79 |  |
| T4 | 5.50 (3.00-8.63) | 76 |  |
| T5 | 6.50 (2.50-9.00) | 53 |  |
| T6 | 5.75 (1.88-11.75) | 38 |  |
| **Moderate type** |  |  | P1 |
| **30 Days** | 4.00 (1.50-7.00) | 297 | 0.000 |
| T1 | 2.00 (1.00-4.50) | 58 | 0.222 |
| T2 | 5.00 (1.50-7.00) | 60 | 0.180 |
| T3 | 5.50 (2.75-8.00) | 59 | 0.001 |
| T4 | 5.00 (2.00-7.00) | 60 | 0.003 |
| T5 | 4.50 (1.88-8.63) | 40 | 0.520 |
| T6 | 4.00 (1.50-7.25) | 20 | 0.573 |
| **Severe type** |  |  | P2 |
| **30 Days** | 7.00 (3.25-13.50) | 79 | 0.012 |
| T1 | 3.50 (1.75-7.00) | 16 | 0.802 |
| T2 | 6.00 (3.88-11.75) | 14 | 0.122 |
| T3 | 9.00 (7.25-15.50) | 14 | 0.179 |
| T4 | 8.25 (6.75-15.38) | 12 | 0.379 |
| T5 | 6.75 (3.75-8.00) | 10 | 0.161 |
| T6 | 5.00 (3.00-14.00) | 13 | 0.095 |
| **Critical type** |  |  | P3 |
| **30 Days** | 12.50 (6.50-16.00) | 37 | 0.000 |
| T1 | 3.00 (2.00-6.50) | 9 | 0.385 |
| T2 | 12.50 (8.00-15.75) | 10 | 0.001 |
| T3 | 15.25 (14.13-17.50) | 6 | 0.000 |
| T4 | 14.00 (12.38-15.00) | 4 | 0.002 |
| T5 | 14.00 (13.25-16.00) | 3 | 0.009 |
| T6 | 14.00 (12.00-16.00) | 5 | 0.010 |

P1: moderate type vs. severe type; P2: severe type vs. critical type; P3: moderate type vs. critical type

**Figure 2:**
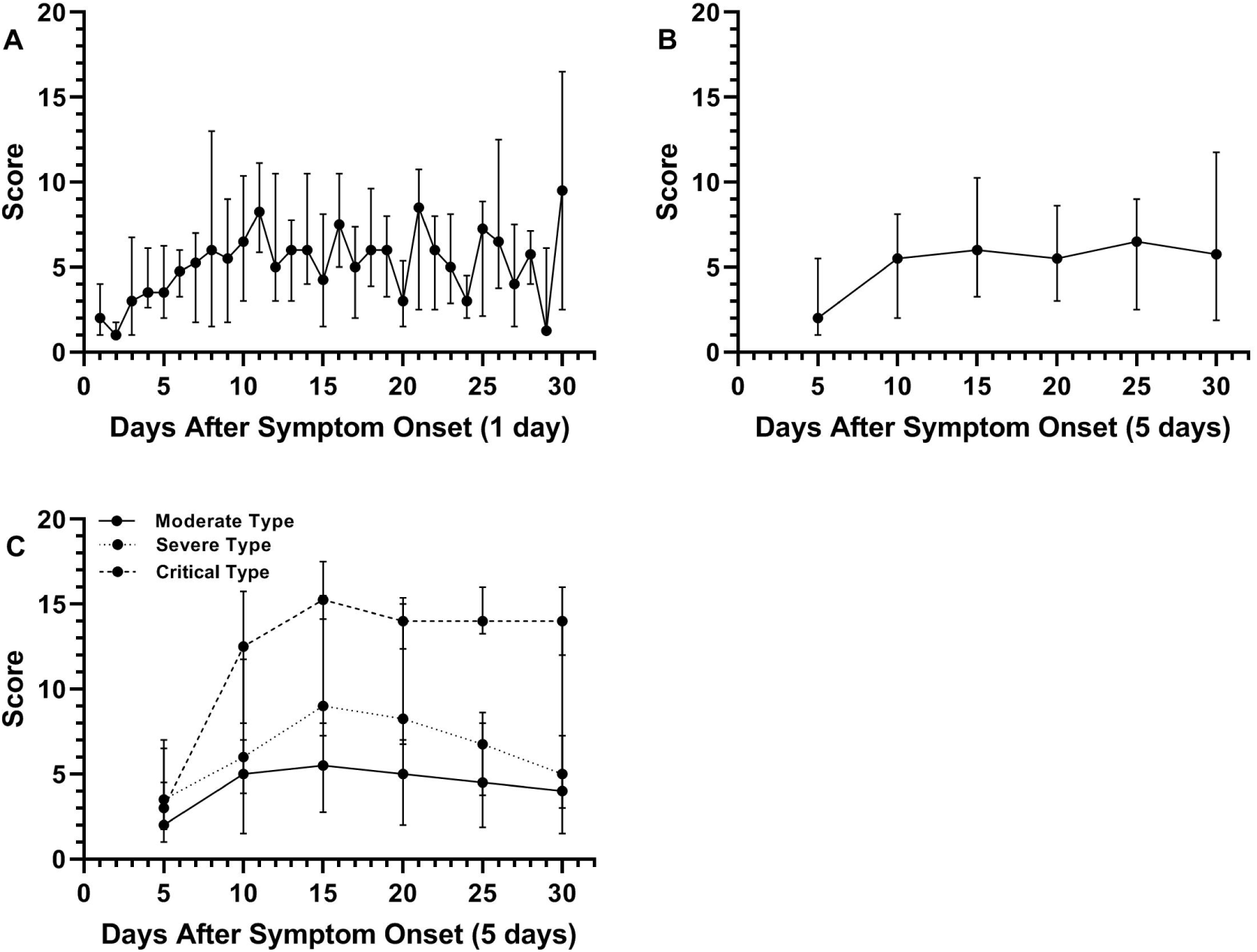
A-B, Temporal changes of the median CT scores in terms of per day (A) and per five days (B) in the whole cohort. C, Temporal changes of the median CT scores in terms of per five days in different clinical types.

The median CT scores of the moderate, severe, and critical type were 4.00 (1.50-7.00), 7.00 (3.25-13.50), and 12.50 (6.50-16.00), respectively (Table 2). The differences among the median scores of the moderate, severe, and critical type were statistically significant (P<0.05). The median scores of severe type were significantly higher than those of moderate type in T3 and T4, and also higher in critical type than moderate type during T2-T6. The changes of the median CT scores in different clinical types were illustrated in Figure 2C.

### Outcomes

Among the 125 patients, 115 patients were discharged from hospital and 10 died. The median CT scores of the survival and death group were 4.50 (1.50-7.00) and 10.25 (3.38-16.00), respectively, with significant difference (P<0.01).

### Categorical regression analysis

We performed a CATREG analysis to assess the association of age, gender, and maximum CT scores during the first 15 days with clinical types of 125 patients (Table 3). The results showed that CT score (P = 0.000) and age (P = 0.000) were significantly, while gender was not significantly (P = 0.184) associated with the clinical type of COVID-19. The age that younger than 60 years old was negatively correlated with clinical type, which was positively correlated with the clinical type when older than 60 years old, the coefficients of 61-70 and >70 years old group were 0.394 and 2.379, respectively. The CT scores of 0.00-11.50, 11.50-16.00, and 16.00-20.00 were separately correlated with the moderate, severe, and critical type. Besides, we classified the patients into different clinical types based on this CATREG model (*Q* _ *level* = 0.559 * *Q* _ *age* + 0.276 * *Q* _ *CT* + 0.077 * *Q* _ *gender*) and compared the results with the actual types (Figure 3). The output accuracy was 69.60%, which was 76.80% when severe and critical type were combined.

**Table 3:**
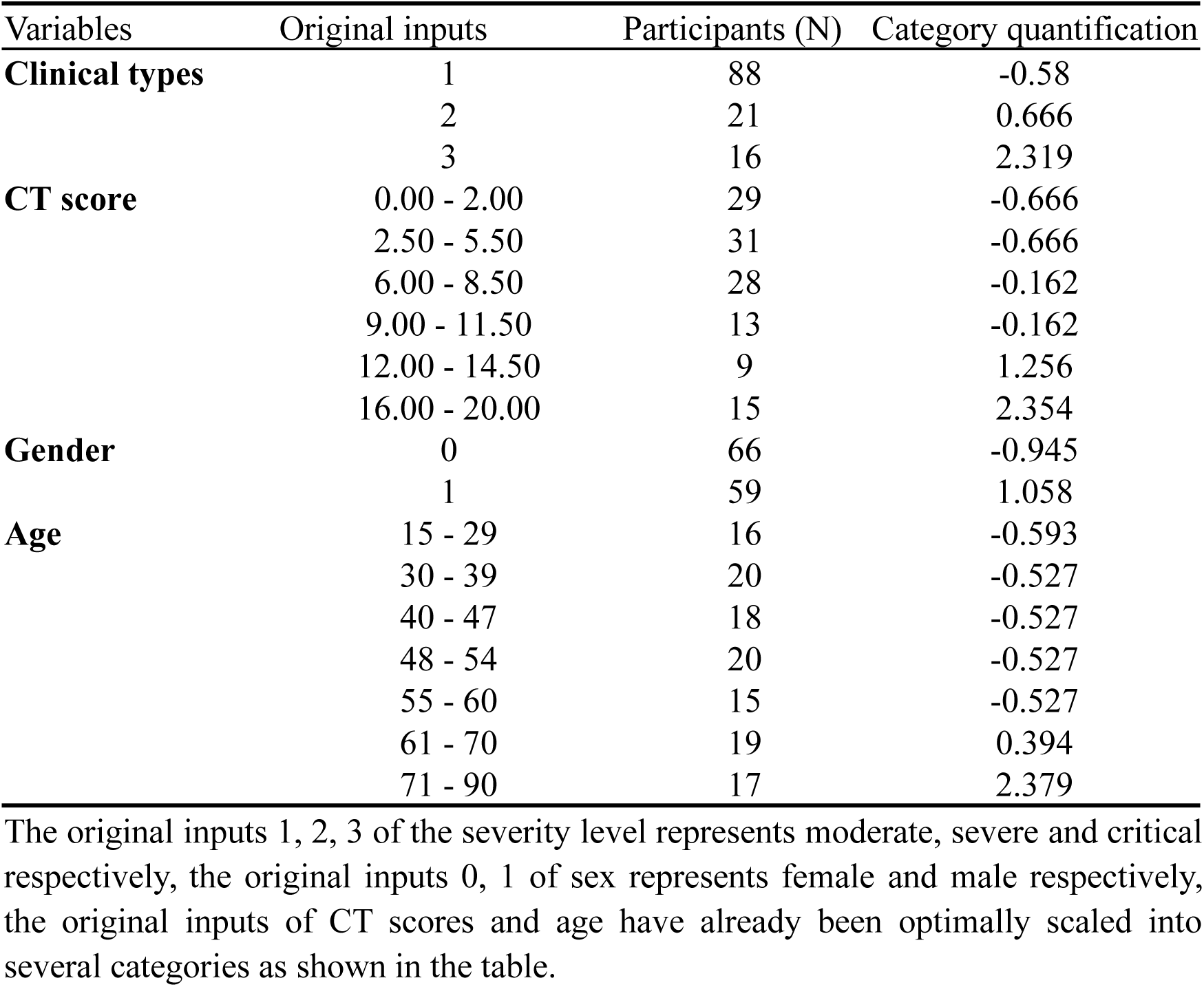
Categorical regression analysis based on optimal scale (CATREG)

| Variables | Original inputs | Participants (N) | Category quantification |
| --- | --- | --- | --- |
| **Clinical types** | 1 | 88 | -0.58 |
|  | 2 | 21 | 0.666 |
|  | 3 | 16 | 2.319 |
| **CT score** | 0.00 - 2.00 | 29 | -0.666 |
|  | 2.50 - 5.50 | 31 | -0.666 |
|  | 6.00 - 8.50 | 28 | -0.162 |
|  | 9.00 - 11.50 | 13 | -0.162 |
|  | 12.00 - 14.50 | 9 | 1.256 |
|  | 16.00 - 20.00 | 15 | 2.354 |
| **Gender** | 0 | 66 | -0.945 |
|  | 1 | 59 | 1.058 |
| **Age** | 15 - 29 | 16 | -0.593 |
|  | 30 - 39 | 20 | -0.527 |
|  | 40 - 47 | 18 | -0.527 |
|  | 48 - 54 | 20 | -0.527 |
|  | 55 - 60 | 15 | -0.527 |
|  | 61 - 70 | 19 | 0.394 |
|  | 71 - 90 | 17 | 2.379 |

The original inputs 1, 2, 3 of the severity level represents moderate, severe and critical respectively, the original inputs 0, 1 of sex represents female and male respectively, the original inputs of CT scores and age have already been optimally scaled into several categories as shown in the table.

**Figure 3:**
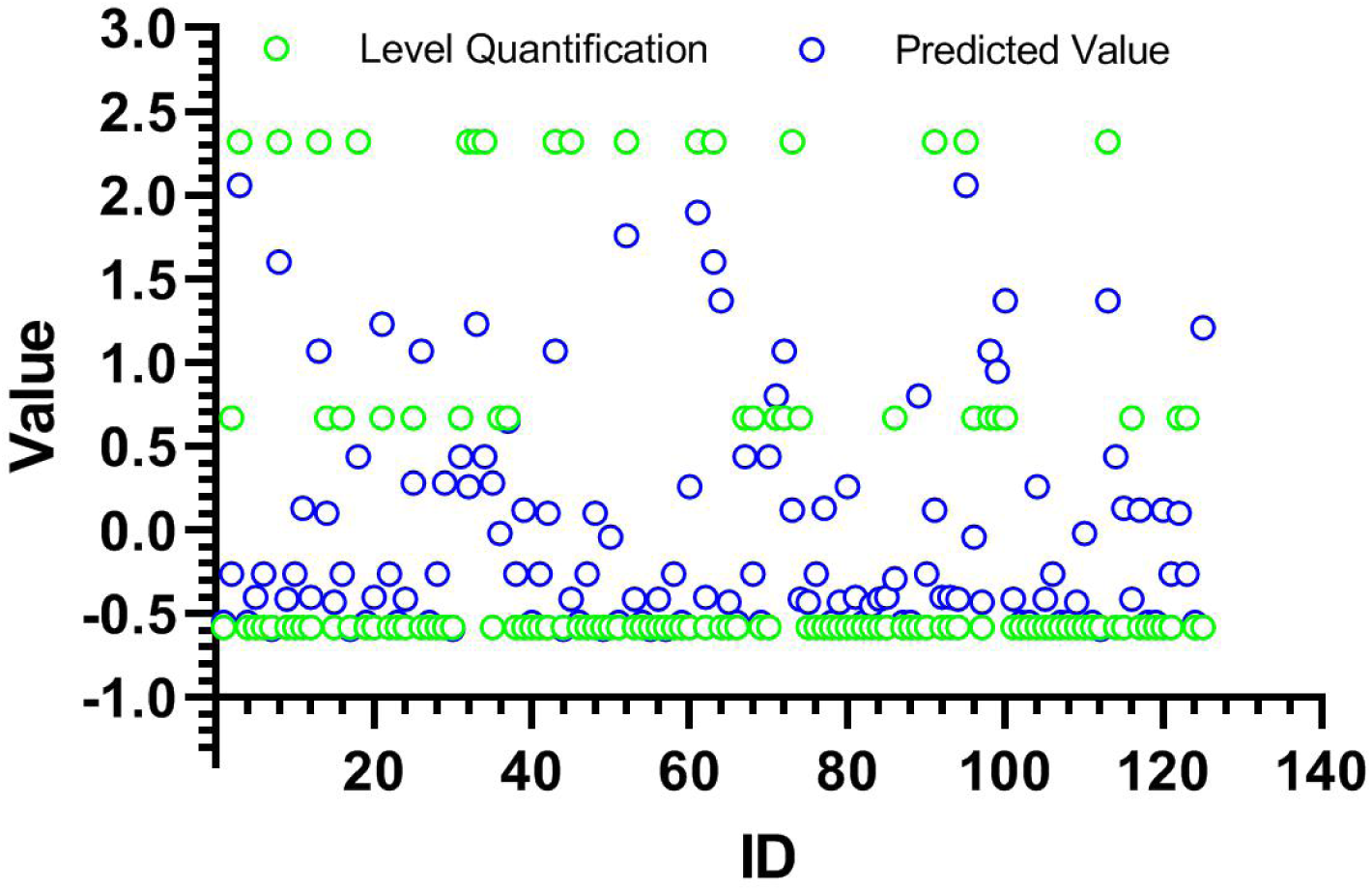
Classification of the clinical types based on CT scores, age, and sex using the CATREG model.

## Discussion

In this study, it was revealed that the durations of the early, progression, consolidation, and dissipation stage of CT images were different from those reported previously^[4, 10, 11]^. The progression and consolidation stages were overlapped in 31.84% (57/179) of the examinations, indicating that the pulmonary exudation and consolidation were coexisting in these CT images. Therefore, the combination of the progression stage and consolidation stage is more consistent with the pathological changes of pulmonary lesions during this period and is helpful to objectively divide stages for CT images of patients with COVID-19. After combination, the appearance time of CT features at progression-consolidation and dissipation stage was longer when the disease was severer. Statistical difference was found between severe and critical type at the combined progression-consolidation stage, suggesting the rationality of our four-stage staging method.

Our data showed that the median CT score on the 1^st^ day was 2.00, suggesting that the pulmonary lesions were already existing when symptom onset. This is consistent with the report that some patients had positive chest CT before symptom onset^[21]^. The peak value of CT scores appeared on the 11^th^ day after symptom onset, and the duration from symptom onset to transforming into critical type was 12.50±5.73 days, indicating that the appearance time of the largest pulmonary lesion is basically consistent with the inception time of critical type. These results also suggest that pulmonary lesions dominate the evolution of COVID-19.

The temporal change trends of median CT scores in terms of every five days showed a plateau during T3-T6. However, the symptoms were relieved and some laboratory test indexes such as CRP, CK, α-HBDH, LDH were decreased in this period (Supplement Figure S1 and Figure S2). This discrepancy between CT scores and illness indicates that the pulmonary lesions are mainly characterized by changes in nature rather than size at the plateau stage, and the CT scores cannot accurately reflect the changes of pulmonary lesions in this period.

The median CT scores of the moderate, severe, and critical type were 4.00, 7.00, and 12.50, respectively, the corresponding peak values were 5.50, 9.00, and 15.25, The median CT score of severe type in T3 was 5.50 higher than T1, demonstrating a trend that the severer the disease, the higher the median CT scores and their peak values; meanwhile when the patients with CT scores >5.50 need to be reexamined with CT every 1-2 days, if their daily scores increase by more than 0.55 or reach about 9 have an increased risk of developing into severe and critical types. Therefore, quantitative analysis of CT images may be a reliable method to evaluate the severity of COVID-19 during 5-15 days.

Besides, the median CT score of the death group was significantly higher than the survival group. The score of the death group may be higher since part of the patients in this group were not convenient to be examined by CT.

According to the regression analysis, the age that younger than 60 years old was negatively correlated with clinical type. For the patients that older than 60 years old, the older the age, the higher the coefficient, and the higher the risk of developing into severe and critical type. Additionally, the CT scores of 0.00-11.50, 11.50-16.00, and 16.00-20.00 were separately correlated with the moderate, severe, and critical type. The clinical type was the foundation of classifying CT images. The standard of typing CT images reflects the relationship between the pulmonary lesion size and lung function, which makes the CT types consistent with the clinical types, and is helpful to analyze the illness and judge the prognosis by CT. The accuracy of this CATREG model was 76.8% when classifying the clinical types into two groups (the moderate type, and the combined severe and critical type), indicating that the CT scores could basically reflect the changes of their pulmonary lesions.

However, the sample size was limited and only the data of patients during hospitalization were studied. Therefore, long-termly follow-up clinical researches are necessary to be carried out with multi-center and more samples.

Overall, the four-stage staging method is rational to describe the temporal distribution of the CT features, and this CT scoring method could quantitatively describe the change of pulmonary lesion size. The results demonstrated that the pulmonary lesions were already existed before symptom onset, the median CT scores were higher with larger rise range and longer declining duration when the disease was severer. The classification standard of CT scores can be used to evaluate the severity of Coronavirus Disease 2019, especially at the stage of pulmonary lesion size rapid enlarged and transforming into severe and critical type.

## Data Availability

Anyone who wants to obtain the original data of this study with reasonable purposes can contact the correspondent author via email.

## Abbreviations

COVID-19: Coronavirus Disease 2019
CT: Computed Tomography
GGO: Ground Glass Opacity
CATREG: Categorical Regression Analysis Based on Optimal Scaling

## Supplement Figure

**Figure S1:**
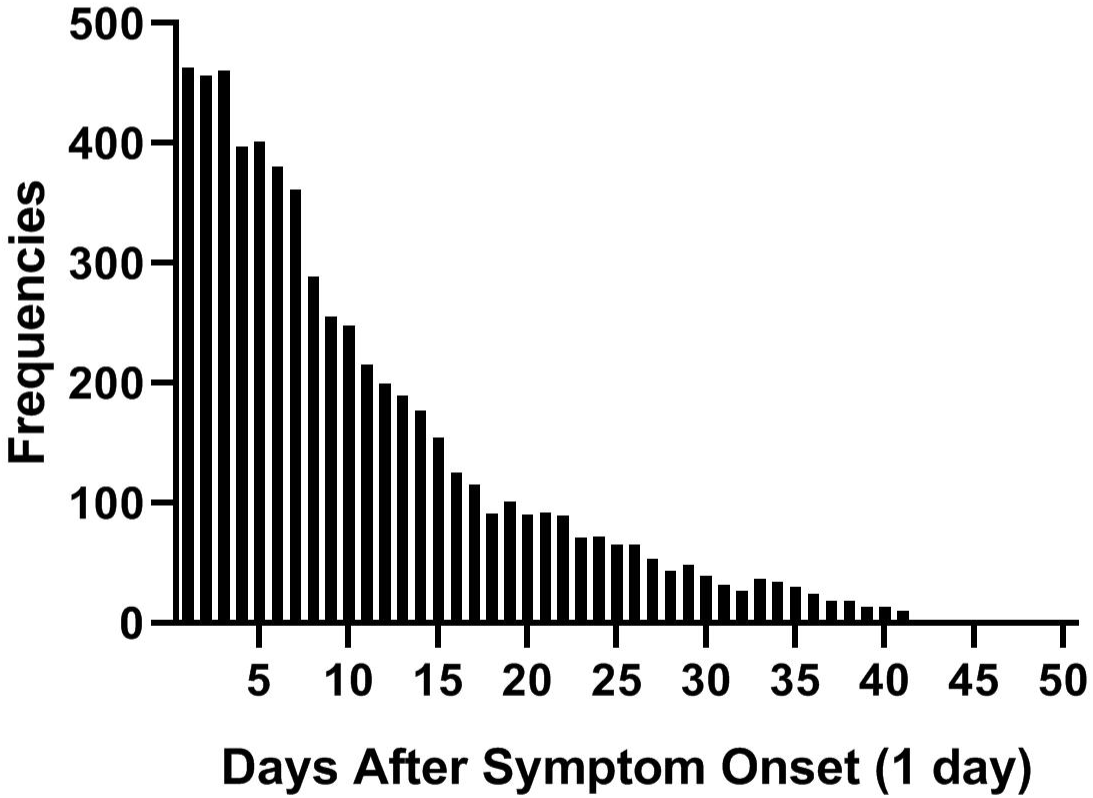
Frequencies of symptoms of the cohort.

**Figure S2:**
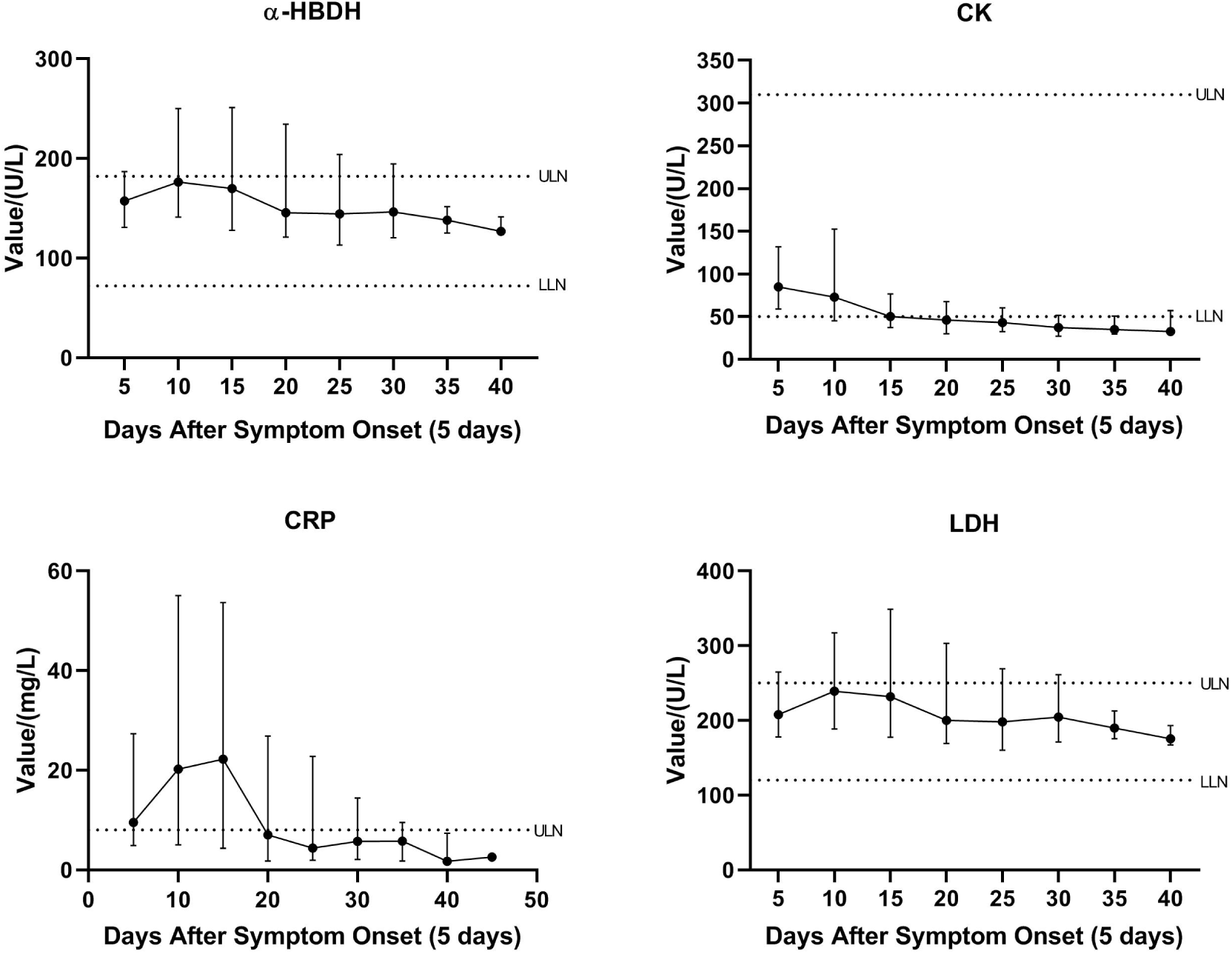
Temporal distribution of the median values of α-HBDH, CK, CRP, and LDH.

